# Absence of relevant QT interval prolongation in not critically ill COVID-19 patients

**DOI:** 10.1101/2020.08.18.20177717

**Authors:** Juan Jiménez-Jáimez, Rosa Macías-Ruiz, Francisco Bermúdez-Jiménez, Ricardo Rubini-Costa, Jessica Ramírez Taboada, Paula Isabel García Flores, Laura Gallo Padilla, Juan Diego Mediavilla García, Concepción Morales García, Sara Moreno Suárez, Celia Fignani Molina, Miguel Álvarez López, Luis Tercedor

## Abstract

**Objectives:** SARS-CoV-2 is a rapidly evolving pandemic causing great morbimortality. Medical therapy with hydroxicloroquine, azitromycin and protease inhibitors is being empirically used, with reported data of QTc interval prolongation. Our aim is to assess QT interval behaviour in a not critically ill and not monitored cohort of patients.

**Design:** We evaluated admitted and ambulatory patients with COVID-19 patients with 12 lead electrocardiogram at 48 hours after treatment initiation. Other clinical and analytical variables were collected. Statistical analysis was performed to assess the magnitude of the QT interval prolongation under treatment and to identify clinical, analytical and electrocardiographic risk markers of QT prolongation independent predictors.

**Results:** We included 219 patients (mean age of 63.6 ± 17.4 years, 48.9% were women and 16.4% were outpatients. The median baseline QTc was 416 ms (IQR 404–433), and after treatment QTc was prolonged to 423 ms (405–438) (P < 0.001), with an average increase of 1.8%. Most of the patients presented a normal QTc under treatment, with only 31 cases (14,1 %) showing a QTc interval > 460 ms, and just one case with QTc > 500 ms. Advanced age, longer QTc basal at the basal ECG and lower potassium levels were independent predictors of QTc interval prolongation.

**Conclusions:** Ambulatory and not critically ill patients with COVID-19 treated with hydroxychloroquine, azithromycin and/or antiretrovirals develop a significant, but not relevant, QT interval prolongation.

**What is already known about this subject?:** Treatment for COVID 19 with empirical drugs hydroxicloroquine, azitromycin and protease inhibitors affects ventricular repolarization by prolonging the QT interval in a variable degree basing upon heterogeneous studies

**What does this study add?:** Not critically ill patients with COVID 19 develop a non relevant QT interval prolongation, leading to a modest arrhythmic risk increase, overall in patients with normal basal ECG

**How might this impact on clinical practice?:** Our findings are significant as ECG monitoring of COVID 19 patients is costly and difficult. Ambulatory and stable patients with normal basal ECG might nor benefit from QT assessment afeter treatment initiation

## BACKGROUND

Since late 2019, a new coronavirus is responsible for the Severe Acute Respiratory Syndrome Coronavirus 2 (SARS-CoV-2) causing a worldwide pandemic.^1^ It has spread rapidly throughout the world with a high mortality rate, especially among older adults with cardiovascular disease.^2^ There is currently no specific preventive or therapeutic strategy approved by the European Medicines Agency (EMA) or the Food and Drug Administration (FDA), so an empiric approach with conventional antimalarial, antibiotic or antiviral drugs are being evaluated for COVID-19. Some of them have been proven to have in-vitro antiviral effects against SARS-CoV-2. Particularly hydroxychloroquine (HCQ) seems to limit the virus entry likely through inhibition of angiotensin-converting enzyme 2 (ACE2) receptors.^3^ Recently, one study has suggested that the concomitant use of azithromycin in combination with HCQ is associated with a more rapid resolution of virus detection than HCQ alone.^4^ Finally, lopinavir/ritonavir is a protease inhibitor largely used in HIV infection, and has in-vitro activity against SARS-CoV-2.^5^ While the optimal strategy to treatment of COVID-19 is uncertain, a combination of these drugs is the accepted worldwide approach in symptomatic SARS-CoV-2 infection.

Beyond their limited evidence for treating COVID-19, the scientific community is aware of adverse effects, in particular the QT interval prolongation. There are sporadic data reporting QT interval prolongation with hydroxychloroquine, when it is used in patients with systemic lupus erythematosus. On the other hand, azithromycin is
a well-recognized ventricular arrhythmia and sudden cardiac death (SCD) aetiology, even in the absence of significative QT interval prolongation.^6–8^ Lastly, lopinavir/ritonavir is listed as a drug with possible risk of torsade de pointes (TdP) at *crediblemeds.org*.

The effect of the combination of one or more of these drugs on cardiac rhythm or repolarization has been recently analysed in studies with heterogeneous sample of COVID-19 patients. Our aim is to study the QT interval behaviour in a selected population of not critically ill patients with COVID-19 treated with hydroxychloroquine, azithromycin and/or antiviral drugs.

## METHODS

### Population and study design

A transversal study was conducted between March 23 and April 24, 2020 in a tertiary referral hospital. Patients presenting to our emergency department (ED) with confirmed diagnosis or high clinical suspicion of SARS-CoV-2 infection and treated whether an inpatient or outpatient management, were identified. Confirmed SARSCoV-2 infection was defined as positive nasopharyngeal polymerase chain reaction (RT-PCR) and/or positive IgM and IgG serological test. High suspicion diagnosis was realized by clinician criteria based on clinical characteristics, chest imaging, analytical parameters and ruling out common infectious pathogens causing pneumonia, if diagnostic tests were undetermined, negative or non-available. Ambulatory and admitted patients with COVID-19 treated with one or more SARS-Cov-2 experimental drugs (according to our hospital protocol, see appendix I) were included. Patients readmitted for SARS-Cov-2 and those admitted to the Intensive Care Unit (ICU) or with mechanical ventilation at the time of baseline and control ECG were excluded. The study was approved by the Local Ethics Committee and verbal informed consent was obtained from participant patients.

### Data collection

Twelve lead ECG was performed in all patients 48h after treatment initiation. In addition, most of the patients had a basal ECG collected before the beginning of medical therapy at the ED admission. Electrocardiographic analysis included heart rhythm and rate, P-QRS-T intervals duration, atrioventricular and intraventricular conduction disturbances and ventricular repolarization in all ECGs. The measurement of the QT interval was preferably carried out on lead II, and V5-V6 if needed, starting at the beginning of the Q wave and using the tangent method described by Postema et al. ^9^ and corrected with the Bazett’s formula. Electrocardiographic assessment was performed by two independent certified cardiac electrophysiologists who were blinded to the patient’s information.

Furthermore, baseline investigations included previous comorbidities, chronic treatments, severity illness at ED, complete blood count, coagulation profile, serum biochemical test (creatine kinase, lactate dehydrogenase, electrolytes, renal and liver function), and complementary treatment during hospitalization with QTc interval prolongation risk were also recorded. Additionally, mortality during admission was analysed.

The primary endpoint of the study was to determine the prevalence of clinically significant QT interval prolongation under medical therapy for COVID-19, especially QTc greater than 500 ms involving a higher arrhythmic risk. Other secondary aims were to detect clinical or analytical independent predictors of QT interval prolongation and quantify the QTc interval increasing from basal ECGs.

### Drug therapy

According to our centre protocol, patients with confirmed or high-suspicion of COVID-19 received a combination of two or more of the following agents: hydroxychloroquine 400 mg BID (loading dose) and 200mg BID during 5 days, azithromycin 500 mg SID (loading dose) and 250mg SID during 5 days, and Lopinavir/Ritonavir 400/100 mg BID or Darunavir/Ritonavir 600/100 mg BID during 7 to 14 days at physician criteria based on severity illness.

### Statistical analysis

Numerical variables are expressed as mean and standard deviation or, for non-normally distributed variables, as median [interquartile range]. Categorical variables are expressed as absolute and relative frequencies. To assess the agreement between the two observers with respect to both, baseline and control QTc parameter measurements, the Bland-Altman graphical method was used, calculated with the interval ± 1.96 times the standard deviation from the mean difference. In addition, the intraclass correlation coefficient has been calculated. The Wilcoxon test was used to compare the baseline and control QTc. A bivariate analysis has been carried out to analyze the variables that are related to the QTc prolongation, establishing 460 ms as the cut-off point. The categorical variables have been contrasted with the Pearson’s chi-square test or Fisher’s exact test in cases where applicability conditions were not met. For the quantitative variables, the Student’s t-test was used for independent samples and the Mann-Whitney test was used for those variables with a non-normal distribution. With the variables in which a p value of <0.20 was obtained, a multivariate logistic regression model was constructed. The variable selection method was by successive steps backwards, removing from the model at each step, those with p <0.10, and using the likelihood ratio method to contrast the appropriate model at each step. For the final model, the odds ratio is calculated along with its 95% confidence interval. The goodness of fit was compared with the Hosmer-Lemeshow test and its predictive capacity was evaluated with the area under the ROC curve.

### Patient and public involvement

There has been no direct public or patient involvement in this research

## RESULTS

### Baseline parameters

Two hundred and nineteen patients with confirmed diagnosis or high suspect of COVID-19 were included, with a mean age of 63.6 (± 17.4 years, 23–98 years), 107 were (48.9%) women. Thirty-six (16.4%) were outpatients. All patients had a control ECG 48 hours after treatment initiation and 165 patients had a baseline ECG at ED admission. At ED admission, 154 patients (93.3%) were on sinus rhythm and 11 on atrial fibrillation (6.7%). One hundred and forty-eight patients (89.7%) had a narrow QRS on admission, 3 had left bundle branch block (1.8%), 9 right bundle branch block(5.5%), 4 bifascicular block (2.4%) and 1 ventricular paced rhythm (0.6%). Patients were divided in two groups according to the treatment: group 1 (single or double therapy) with 105 patients (47.9%) who received hydroxychloroquine and/or azithromycin and group 2 (triple therapy) with 114 patients (52.1%) who received hydroxychloroquine, azithromycin and protease inhibitors (lopinavir-ritonavir or darunavir-ritonavir). Only one patient (0.5%) did not received azithromycin and 9 did not received hydroxychloroquine (4.1%). Baseline characteristics according to treatment are shown in table 1; patients with triple therapy had a worse clinical presentation during admission, with more fever, more multilobar pneumonia and worse analytics parameters.

**Table 1.** Baseline and clinical characteristics depending of COVID-19 treatment administered.

|  | Total<br>(N=219) | Hydroxychloroquine<br>and/or azithromycin<br>(N=105)<sup>1</sup> | Hydroxychloroquine,<br>azithromycin and<br>antiretroviral<br>(N=114) | *P*<br>value |
| --- | --- | --- | --- | --- |
| **Baseline characteristics–no. (%)** |  |  |  |  |
| Age, mean (SD)–yr | 63.6 (17.4) | 67.1 (19.3) | 60.3 (14.7) | .002 |
| Male sex | 111 (50.7) | 46 (43.8) | 66 (57.9) | .037 |
| AHT | 109 (49.8) | 55 (52.4) | 54 (47.4) | .459 |
| Structural Heart<br>Disease<sup>2</sup> | 33 (15.1) | 21 (20.0) | 12 (10.5) | .050 |
| Obesity | 19 (8.7) | 8 (7.6) | 11 (9.6) | .594 |
| ACEI/ARB | 83 (37.9) | 39 (37.1) | 44 (38.6) | .825 |
| Beta blocker | 39 (17.8) | 22 (21.0) | 17 (14.9) | .243 |
| Antidepressant | 36 (16.4) | 26 (24.8) | 10 (8.8) | .001 |
| **Clinical features–no. (%)** |  |  |  |  |
| Fever–no. (%) | 145 (66.2) | 56 (53.3) | 89 (78.1) | <.001 |
| Multilobar<br>Pneumonia–no. (%) | 135 (61.6) | 47 (44.8) | 88 (77.2) | <.001 |
| Ferritin–median<br>[IQR], ng/mL | 573.0<br>[205.5-1275.0] | 330.0 [170.0-514.0] | 1040.0 [394.3-2215.6] | <.001 |
| Hs Troponin I,<br>median [IQR],<br>pg/ml | 6.1 [2.2-20.0] | 11.0 [4.0-31.0] | 4.7 [2.1-10.6] | .005 |
| DD–median [IQR],<br>mg/L | 0.9 [0.5-2.1] | 1.44 [0.65-3.9] | 0.95 [0.6-1.5] | .203 |
| LDH–median<br>[IQR], IU/L | 364.0<br>[271.0-458.0] | 308.0 [268.0-405.0] | 391.5 [347.5-521.3] | <.001 |
| CRP–median<br>[IQR], mg/L | 73.0 [20.0-156.0] | 52.0 [22.5-172.0] | 84.0 [39.5-177.5] | <.001 |
| K<sup>+</sup>–mean (SD),<br>mEq/L | 4.0 [3.8-4.3] | 4.0 [3.8-4.4] | 4.0 [3.8-4.3] | .494 |
| Ca<sup>2+</sup>–mean (SD),<br>mg/dL | 9.2 [8.8-9.6] | 9.2 [8.6-9.6] | 9.2 [8.9-9.5] | .374 |
| Tocilizumab–no.<br>(%) | 11 (5.0) | 0 (0.0) | 11 (9.6) | .001 |
| Antidepressant–no.<br>(%) | 27 (12.3) | 19 (18.1) | 8 (7.0) | .013 |
| Neuroleptic–no.<br>(%) | 19 (8.7) | 14 (13.3) | 5 (4.4) | .019 |
| Exitus–no. (%)*** | 15 (8.2) | 10 (14.5) | 5 (4.4) | .016 |

<sup>1</sup> Note that 90.5% (n=95) of the patients received both drugs, azithromycin and hydroxychloroquine.
<sup>2</sup> Considered: ischemic, hypertensive, dilated, hypertrophic and valvular cardiomyopathy. <sup>3</sup>Inpatient mortality (n=183). (ACEI, angiotensin converting enzyme inhibitor; AHT, arterial hypertension ARB, angiotensin receptor blocker; CRP, C-reactive protein; DD, D-dimer; ECG, electrocardiogram; Hs, high sensitive; IQR, interquartile range; LDH, lactate dehydrogenase; SD, standard deviation)

### QTc interval Analysis

Correlation between both ECG observers was excellent basing upon the result of the Blant Altman test (Suplementary figure 1) with an intraclass coefficient of 0.903.

The spectrum of QTc distribution after treatment is shown in figure 1, with only 4 patients with QTc > 500 ms, most of them cases with wide QRS due to intraventricular conduction disturbances or paced rhythm; one case with narrow QRS complex developed a QTc interval > 500 ms. Only 14,1 % of the cases (n=31) presented a QTc interval of more than 460 ms after treatment and no patient presented *torsade de pointes* or arrhythmic death during admission. Table and figure 2 show a bivariate model to identify variables associated with the presence of a QTc> 460 ms. There were no differences in QTc between treatment groups 1 and 2.

**Figure 1.**
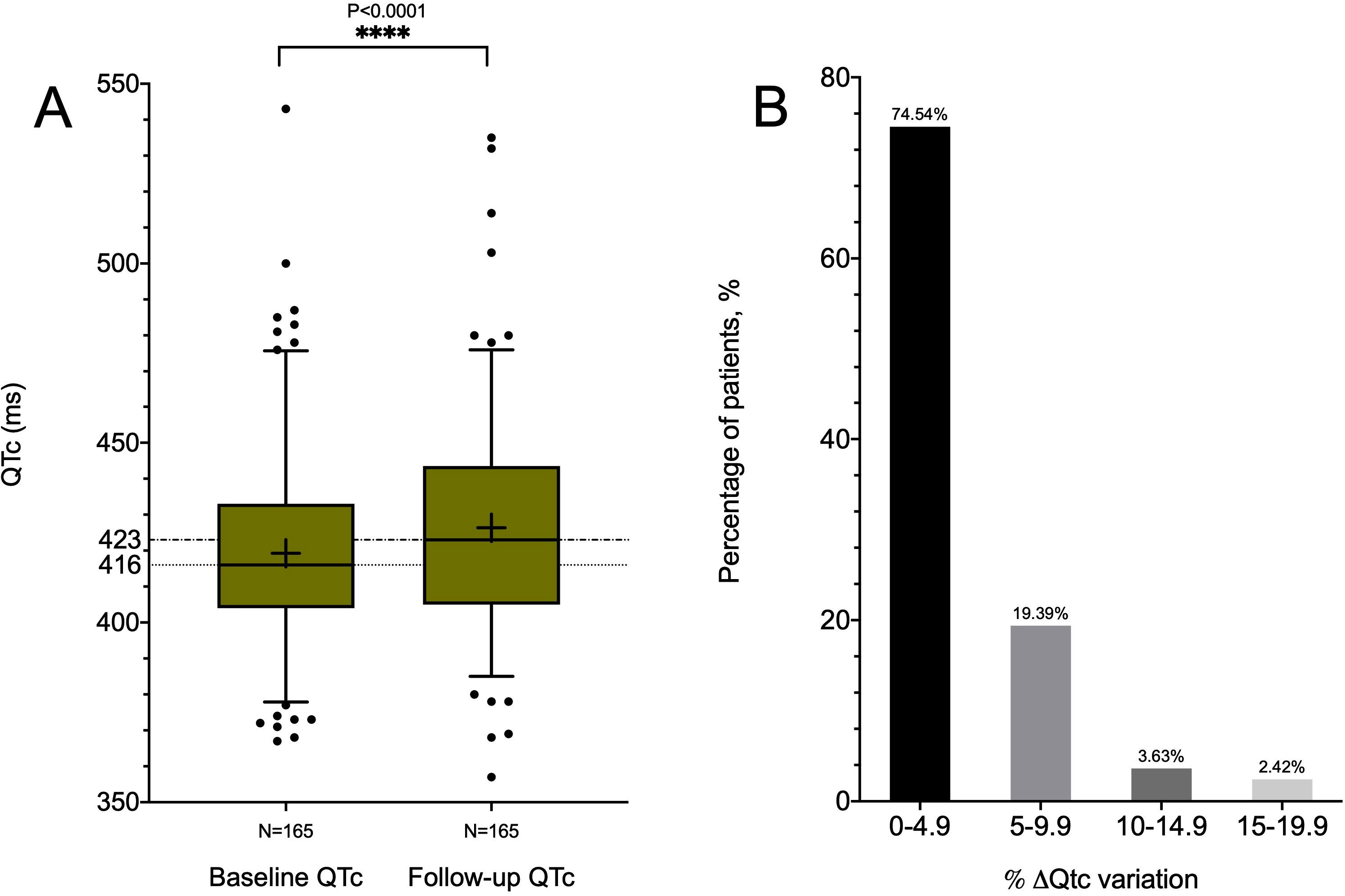
Distribution of patients according to the QTc interval during treatment, at 10 ms intervals. Only 2.9% (n=6) of patients presented a QTc of more than 480ms. Among the 4 patients presenting a QTc interval of ≥500 ms, 3 showed a wide QRS due to intraventricular conduction defects (left bundle branch block or right bundle branch block)

**Figure 2.**
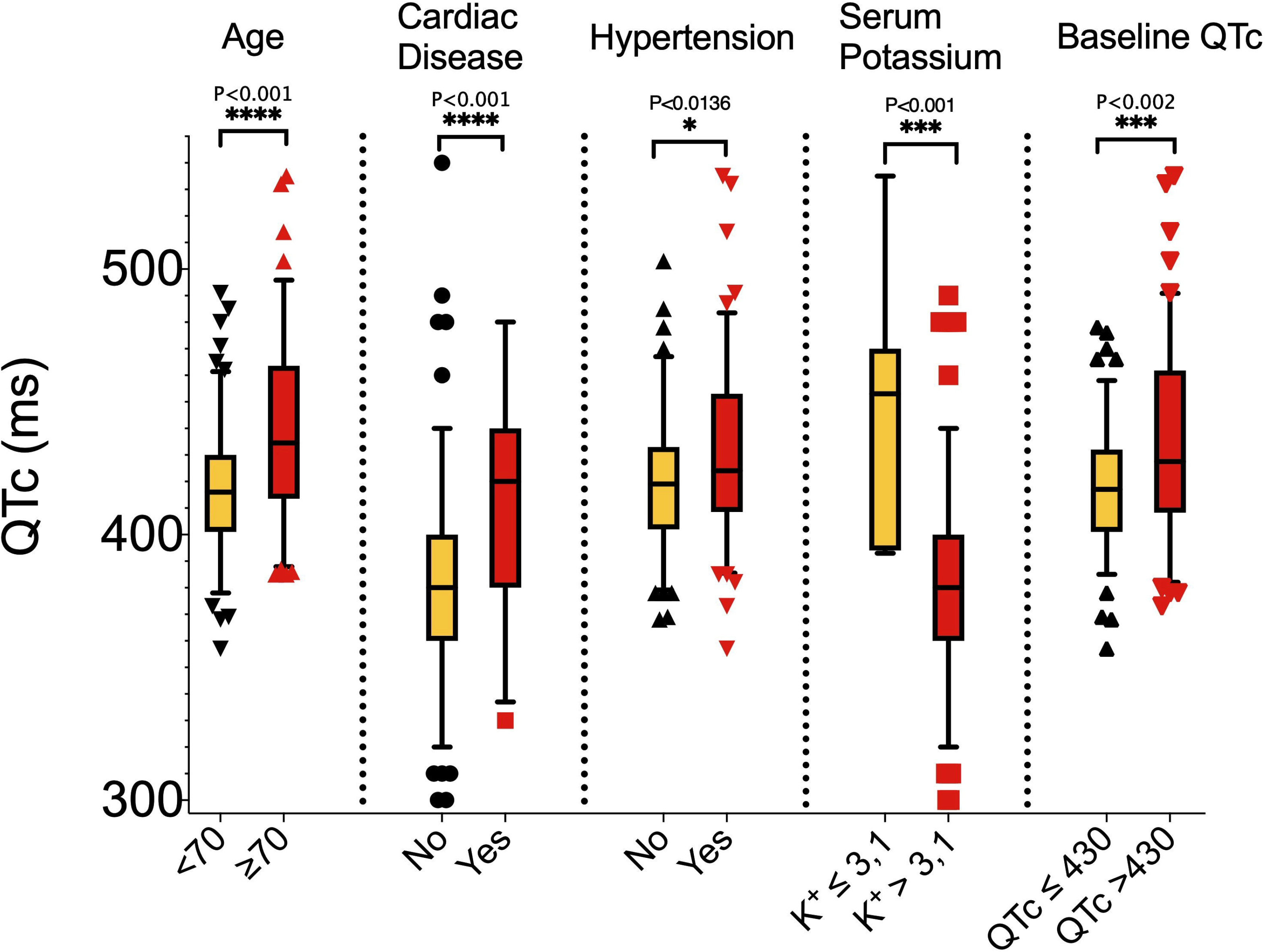
Representation of QTc interval of relevant variables associated with the presence of a QTc> 460 ms (cardiac disease, age, hypertension and the previous treatment with antidepressants).

**Table 2.** Baseline, ECG and clinical characteristics of patients according to the QTc interval at ≥ 48h from treatment initiation.

|  | QTc ≤460 ms<br>(n=188)<sup>1</sup> | QTc >460 ms<br>(n=31)<sup>1</sup> | P value |
| --- | --- | --- | --- |
| Baseline characteristics–no. (%) |  |  |  |
| Age–mean (SD), yr | 61.3 (17.0) | 77.3 (2.9) | <.001 |
| Male sex | 99 (52.7) | 13 (41.9) | 0.268 |
| Outpatient management | 35 (18.6) | 1 (3.2) | .032 |
| AHT | 87 (46.3) | 22 (71.0) | .011 |
| Structural Heart Disease | 20 (10.6) | 13 (41.9) | <.001 |
| Obesity | 18 (9.6) | 1 (3.2) | .487 |
| ACEI/ARB | 66 (35.1) | 17 (54.8) | .046 |
| Beta blocker | 31 (16.5) | 5 (25.8) | .209 |
| Antidepressant | 24 (12.8) | 12 (38.7) | <.001 |
| ECG at baseline–median (IQR), |  |  |  |
| PR interval, ms | 150 [140-160] | 150 [140-180] | 0.915 |
| QRS interval, ms | 90 [80-100] | 100 [90-122] | <.001 |
| QT interval, ms | 360 [330-370] | 360 [340-400] | 0.125 |
| QTc interval, ms | 414 [400-427] | 446 [433-476] | <.001 |
| Heart Rate, bpm | 81 [72-99] | 94 [75-101] | 0.113 |
| Clinical features–median (IQR) |  |  |  |
| Length of stay, d | 7.5 [5.0-11.0] | 7.0 [5.0-9.8] | .499 |
| Fever–no. (%) | 126 (67.0) | 19 (61.3) | .532 |
| Multilobar Pneumonia–no. (%) | 112 (59.6) | 23 (74.2) | .121 |
| Ferritin–ng/mL | 571.0 [196.8-1337.3] | 642.0 [258.0-1119.0] | .771 |
| Hs Troponin I–pg/ml | 5.0 [2.0-13.0] | 22.4 [9.4-69.0] | <.001 |
| DD–mg/L | 0.9 [0.5-1.9] | 1.4 [0.8-13.5] | .008 |
| LDH–IU/L | 361.5 [262.3-457.5] | 392.0 [301.0-505.0] | .175 |
| CRP–mg/L | 67.0 [18.1-146.0] | 78.0 [37.0-209.0] | .084 |
| Na+–mEq/L<sup>2</sup> | 139.0 [136.0-141.0] | 138.5 [136.3-141.0] | .649 |
| K+–mEq/L<sup>2</sup> | 4.1 [3.8-4.4] | 3.7 [3.4-4.3] | .087 |
| Ca<sup>2+</sup>–mg/dL<sup>2</sup> | 9 [8.6-9.4] | 8.5 [8.3-9.0] | .009 |
| Tocilizumab–no. (%) | 9 (4.8) | 2 (6.5) | .658 |
| Antidepressant–no. (%) | 16 (8.5) | 11 (35.5) | <.001 |
| Neuroleptic–no. (%) | 13 (6.9) | 6 (19.4) | .035 |
| Treatment discontinuation–no. (%)<sup>3</sup> | 6 (3.2) | 8 (25.8) | <.001 |
| Exitus–no. (%) | 10 (5.3) | 5 (16.1) | .027 |

<sup>1</sup>Both groups were created using a QTc cut-off of 460ms in the ECG at ≥48h from treatment initiation. <sup>2</sup>Plasma ions were analyzed at the time the control ECG was performed. <sup>3</sup>Treatment discontinuation because of findings in the ECG at ≥48h from treatment initiation. (ACEI, angiotensin converting enzyme inhibitor; AHT, arterial hypertension ARB, angiotensin receptor blocker; CRP, C-reactive protein; DD, D-dimer; ECG, electrocardiogram; Hs, high sensitive; IQR, interquartile range; LDH, lactate dehydrogenase; SD, standard deviation)

The median baseline QTc was 416 ms (IQR 404–433), and after treatment QTc was prolonged to 423 ms (405–438) (P < 0.001) (figure 3, panel A). An increase in QTc between 0 and 5% was observed in most of the patients, with an average increase of 1.8% (–1.3 to 5.6%) (figure 3, panel B); importantly, no patient showed an increase greater than 25%.

**Figure 3.**
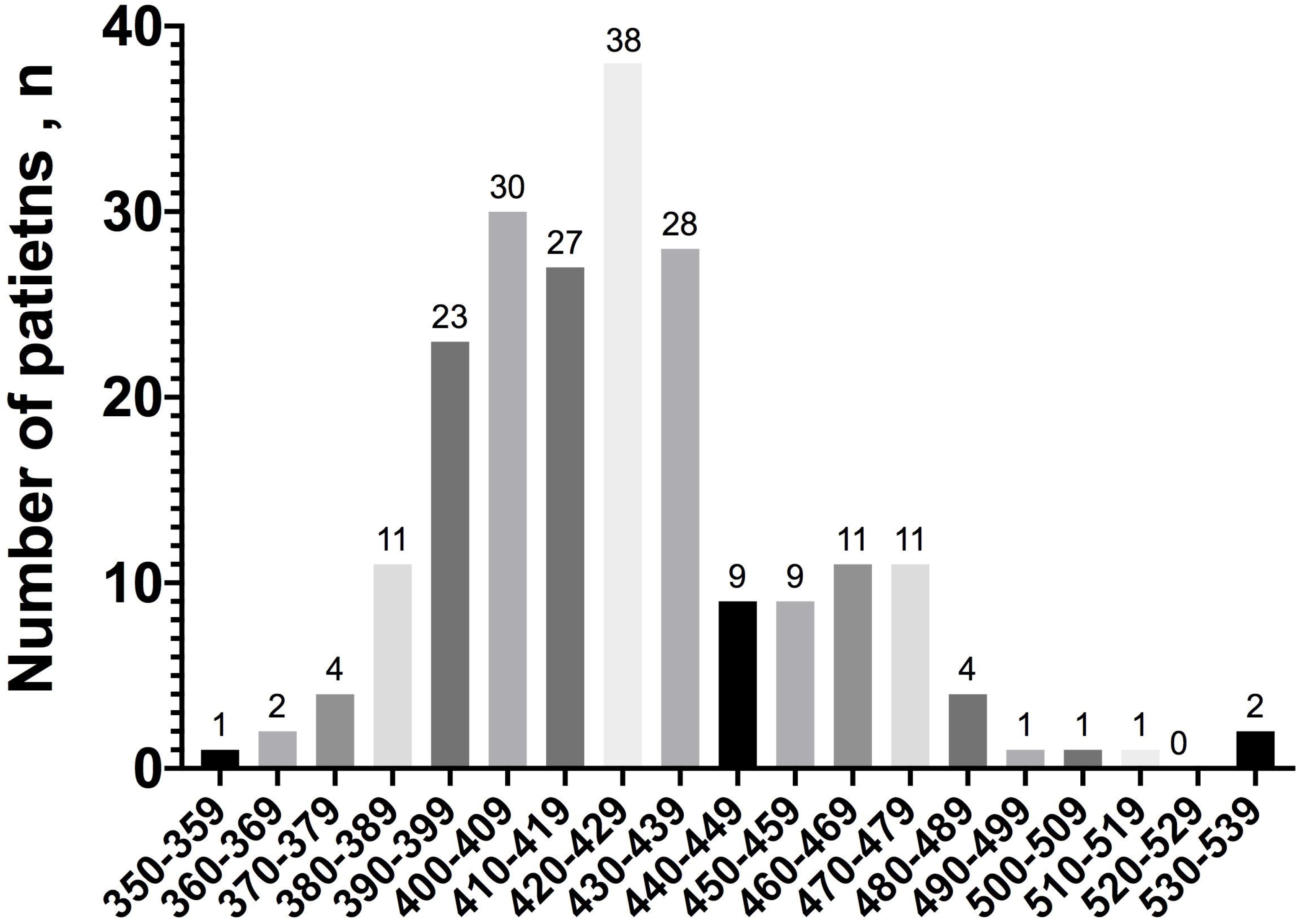
Panel A. Representation of QTc interval at baseline (at treatment initiation) and at follow-up (at least 48h on treatment). The median of the baseline QTc was 416 (404–433) ms; after treatment QTc was prolonged to 423 (405–438) ms. **Panel B. Distribution of patients according to the percentage increase of the QT interval with respect to the basal**. The vast majority of patients presented a mild (0–9.9%) raise in QTc during treatment.

In the multivariate model (table 3) age was a risk factor, with an OR 1.05 [1,009–1,095]. For each year of age, the risk of prolonged QTc increased by 5%. The baseline QTc is a risk factor, with an OR 1.06 [1,009–1,095]. For each ms of increasing in baseline QTc interval, the risk of prolonged QTc increased by 5%. Basal potassium acts as a protective factor. For each unit that increases basal potassium, the risk of prolonged QTc is 3.29 times lower (1/0.3039). Predictive ability of the model was very good, with an area under the ROC curve of 0.9130 (suplementary figure 2).

**Table 3.** Multivariate analysis. (CI, confidence interval; OR, odds ratio. Sig, statistical significance)

|  | Sig. | OR | OR 95% CI |  |
| --- | --- | --- | --- | --- |
|  |  |  | Lower | Upper |
| Age | .0015 | 1.051 | 1.009 | 1.094 |
| K+ at baseline | .043 | 0.30 | 0.095 | 0.964 |
| QT interval at baseline | <.001 | 1.061 | 1.032 | 1.091 |

### Follow-up

Fourteen patients discontinued treatment (6.4%), being in 11 patients (5%) due to QTc prolongation. The median hospital stay was 7 (5–11) days. After admission, 5 patients required intensive care, and only one of them in needing of orotracheal intubation. Fifteen patients died (6.8%) and two of the outpatients required admission during follow-up due to worsening symptoms.

## DISCUSSION

Scientific community is facing against one of the hardest epidemiological crisis in the last century, and safety regarding medical therapies used against SARS-Cov-2 infection seems critical to avoid iatrogenia. Indeed, significant concern about ventricular arrhythmias and SCD in COVID-19 population have emerged, since previous data about QT interval prolongation are reported with the use of hydroxychloroquine and azithromycin, which added to the antiretroviral drugs have been defined as “the corona cocktail”^10^. In this direction, a recent report from the Italian group in Lombardia has warned about the important increase in out-of-hospital cardiac arrests, suggesting a hypothetical link between SARS-CoV-2 infection and this high rate of ventricular arrhythmias^11^. One of the connections might be the QTc prolongation and torsade de pointes induced by drugs (DI-TdP) in ambulatory and admitted, but not ECG-monitored, medically treated patients with COVID-19. Our study focuses on this specific group of patients that, due to a not so serious SARS-CoV-2 disease stage, are not admitted to the ICU, or they even are discharged without admission to hospital. The lack of ECG monitoring would markedly reduce the survival chances of these patients in case of DI-TdP. Our data are strongly reassuring about the use of this combination of drugs in this specific cohort of patients, as we have not detected a clinically relevant QTc interval prolongation attributable to the treatment. Interestingly, our serie is the first, as far as we know, to test the safety of the combination of the three drugs, including antiretrovirals, against COVID-19.

There are some recent reports describing a high percentage of patients developing significant QT interval prolongation (more than 500 ms) in patients with COVID-19, albeit in heterogeneous cohort of patients^12–14^. A recent report from the New York group found a significant QTc interval prolongation in a substantial proportion of patients, being more pronounced in patients with congestive heart failure, on amiodarone or with higher basal QTc intervals^12^. They even present a case of DI-Tdp successfully defibrillated. However, the report of TdP cases in COVID-19 patients is scarce and it likely occurs in ECG-monitored patients admitted to the ICU. Strikingly, their median basal QTc interval is quite long, about 440 ms, which is the upper limit for healthy population; something similar was observed in other works with a basal median QTc of 455 ms^13,14^. This fact suggests the presence of concomitant factors predisposing this extreme QTc interval prolongation response.^14^ Our data suggest a modest QTc interval prolongation, with just one case, in the absence of LBBB or ventricular paced QRS, greater than 500 ms.

There may be several reasons for this different observation. First of all, ours is a homogenous cohort of patients excluding severely ill patients in needing of mechanical ventilation nor central nervous system depressants drugs; basal QTc in our serie is similar to the one observed in general population^15^. Previous studies with extremely prolonged basal QT intervals may not reflect the patient’s real-world baseline repolarization status. This data have been replicated recently by the group from Yale^16^, that describe a significantly prolonged QTc interval after treatment among patients admitted to ICU after reviewing more than two thousands ECGs of patients with COVID-19; on the other hand, previous studies focus mostly on American population^12–15^, with lack of large series in other regions. A single European work showed a significantly prolonged QT interval in more than 20 % of the sample^17^, but in this case patients were treated with chloroquine, which is known to affect QT interval in a greater degree than HCQ. In fact, chloroquine has even been tested in a randomized clinical trial for patients with COVID-19, showing significant QT interval prolongation, although in higher doses than the conventional used^18^. Another European cohort in a very limited sample of patients, most of them admitted to the ICU and intubated, showed a significant proportion of patients with QTc interval greater than 500 ms^19^. These data differ with ours, although basal population and treatment approach is different too. A recent multicentric registry has shown an increased risk of arrhythmic risk in patients taking antimalarial +/− macrolide, but with no clear relation with an underlying QT interval prolongation^20^.

It is well recognized that genetic background is the key point to develop QTc interval prolongation and DI-TdP. Some genetic polymorphisms are common in specific geographical areas and may predispose, under environmental circumstances such as drugs or electrolyte disturbances, to prolong the QT interval. Particularly, some polymorphisms in SCN5A are present in a high proportion of American population of African-descent, such as p.Ser1103Tyr; this variant is absent in Europeans and has been proven to increase the late/persistent sodium current, with a basal modest proarrhyhtmic effect, but an enhanced risk of DI-TdP under clinical scenarios that reduce the repolarization reserve such as hypoxia, acidosis or adverse clinical status with advanced age,^10,21–27^. This is something we have also observed in our serie, as advanced age was an independent predictor of QT interval prolongation. However, in the absence of a proarrhythmic genetic variant, these unfavourable conditions seem to be not enough to damage the repolarization reserve and are not able to create the perfect substrate for the appearance of ventricular arrhythmias and SCD. This fact might be contributing to the observed difference between our serie and the ones by the Americans.

Taking our data into consideration, our hypothesis to explain the higher rate of out of hospital cardiac arrests described by some groups^11^ does not support the DI-TdP etiology. Recent studies have shown a worrisome decline in the number of patients treated with primary coronary intervention for acute coronary syndrome during the pandemia^28,29^. Fear of patients to be admitted to health centers as well as the strict public health laws may have contributed to this phenomenon, in a higher grade than the sporadic cases of DI-TdP in patients with COVID-19.

## CONCLUSIONS

Ambulatory and not critically ill patients with COVID-19 treated with hydroxychloroquine, azithromycin and/or antiretrovirals develop a significant, but not relevant, QT interval prolongation. Advanced age, prolonged baseline QTc and low basal potassium levels are independent markers for this prolongation. Although caution is needed to identify basal risk predictors, these data are reassuring for this specific population

## LIMITATIONS

First, due to the centre’s treatment protocol all patients received the medication and a control group of untreated positive COVID-19 patients was not available. Second, the absence of baseline ECG in some patients and limited follow-up may underestimate the real incidence of high risk QTc prolongation.. Finally, considering the daily update on the disease, we may have not reflected on some relevant clinical variables that could have confounded clinical conclusions.

## Data Availability

Available at DRYAD public repository

## ACKNOWLEDGMENTS

We are grateful to our colleagues who contributed collecting the ECGs from the COVID-19 patients, in such a difficult context, as well to nurses and auxiliary and administrative personal that have taken part in this project in different ways. Particularly thankful we are to Manuel Molina, Pablo Sánchez Millán, Juan Emilio Alcalá López, Laura Pertejo Manzano, Vicente Alcalde Martínez, Ricardo Rivera López, Joaquín Sánchez Gila, Guillermo Gutiérrez Ballesteros, Lorena González Camacho, Emilio Quintero, Rocío Parrilla Linares, Miguel Morales García, Jesús Peña, Antonio Arriaga, Navarrete, José Manuel Díaz López, Ana Pardo and Laura Lamarca and Rosario García Constan de la Revilla, Alicia Almagro Romero, Eva Cabrera Borrego, Lucía Torres Quintero,Rocío Parrilla Linares, Miguel Morales García, Jesús Peña, Antonio Arriaga,Jose Sánchez Moreno, Isabel Merino, Jesús López Muñoz, Elena Sola García, Nuria Navarrete, José Manuel Díaz López, Ana Pardo and Laura Lamarca and Rosario García and Juan Jiménez Alonso.

## a) Contributorship Statement

All the authors has contributed to the planning, conduct, or reporting of the work described in the article; Dr Juan Jiménez Jáimez is responsible for the overall content as guarantor(s).

## b)Funding Statement

nothing to declare

## c) Competing interests

nothing to declare

---

**Figure S1.** Blant-Altman test showing a high grade of concordance between both observers

**Supplementary material:** COVID-19 internal management protocol.

**Figure S2.** Predictive ability of the multivariate analysis, area under the ROC curve.

## SUMMARY BOX

What is already known on this topic

- The absence of evidence-based treatment during the SARS-CoV-2 pandemic has led to the worldwide use of drugs such as hydroxychloroquine, azithromycin and antiretrovirals on an empirical basis.
- Recent studies have warned of the risk of QT prolongation-related arrhythmic adverse events in COVID-19 patients treated with hydroxychloroquine and azithromycin, usually in severe or critical situations.
- This evidence stems from observational studies comprising patients usually in critical condition with an increased intrinsic risk of major arrhythmic events.

What is new and what are the clinical implications?

- For the first time, we describe the arrhythmogenic impact of hydroxychloroquine plus azithromycin in a cohort of non-severe ill patients, including outpatient management.
- Patients presenting COVID-19 infection with mild to moderate affection are not at risk of severe cardiac rhythm disturbances due to combined hydroxychloroquine and azithromycin treatment
- Patients with advanced age, prolonged baseline QTc or low basal potassium levels are at higher risk of QT interval prolongation, and treatment should be administered with caution

